# Associations of circulating T-cell subsets in carotid artery stiffness: the Multi-Ethnic Study of Atherosclerosis

**DOI:** 10.1101/2024.07.29.24311196

**Authors:** Theodore M. DeConne, Petra Buzkova, Ryan Pewowaruk, Joseph A. Delaney, Bruce M. Psaty, Russell P. Tracy, Margaret F. Doyle, Colleen M. Sitlani, Alan L. Landay, Sally A. Huber, Timothy M. Hughes, Alain G. Bertoni, Adam D. Gepner, Nels C. Olson, Jingzhong Ding

## Abstract

**Background:** Arterial stiffness measured by total pulse wave velocity (T-PWV) is associated with increased risk of multiple age-related diseases. T-PWV can be described by structural (S-PWV) and load-dependent (LD-PWV) arterial stiffening. T-cells have been associated with arterial remodeling, blood pressure, and arterial stiffness in humans and animals; however, it is unknown whether T-cells are related to S-PWV or LD-PWV. Therefore, we evaluated the cross-sectional associations of peripheral T-cell subpopulations with T-PWV, S-PWV, and LD-PWV stiffness.

**Methods:** Peripheral blood T-cells were characterized using flow cytometry and the carotid artery was measured using B-mode ultrasound to calculate T-PWV at the baseline examination in a subset of the Multi-Ethnic Study of Atherosclerosis (MESA, n=1,984). A participant-specific exponential model was used to calculate S-PWV and LD-PWV based on elastic modulus and blood pressure gradients. The associations between five primary (p-significance<0.01) and twenty-five exploratory (p-significance<0.05) immune cell subpopulations, per 1-SD increment, and arterial stiffness measures were assessed using adjusted, linear regressions.

**Results:** For the primary analysis, higher CD4^+^CD28^-^CD57^+^ T-cells were associated with higher LD-PWV (β=0.04 m/s, p<0.01) after adjusting for co-variates. For the exploratory analysis, T-cell subpopulations that commonly shift with aging towards memory and differentiated/immunosenescent phenotypes were associated with greater T-PWV, S-PWV, and LD-PWV after adjusting for co-variates.

**Conclusions:** In this cross-sectional study, several T-cell subpopulations commonly associated with aging were related with measures of arterial stiffness. Longitudinal studies that examine changes in T-cell subpopulations and measures of arterial stiffness are warranted.

## INTRODUCTION

Stiffening of the large elastic arteries occurs with aging, and is a risk factor for age-related cerebrovascular dysfunction and neurodegenerative disorders, chronic kidney disease, diabetes, cardiovascular disease (CVD), stroke, and death^1–8^. Arterial stiffness occurs via multiple mechanisms, including increased pulsatile load from elevated blood pressure (load-dependent), and structural changes to the vasculature, such as elastin degradation, and collagen deposition (structural stiffening)^9,10^. New methods in measuring arterial stiffness allow for differentiation of the load-dependent (LD-PWV) component from the structural (S-PWV) arterial stiffness component^10^.

Adaptive T-lymphocytes (T-cells) have been associated with arterial stiffness and its associated components (e.g., arterial remodeling and blood pressure)^11–17^. For example, CD4^+^CD45RA^+^ T-cells were associated with lower common carotid artery intima-media thickness (CCA IMT) and higher proportions CD4^+^CD45RO^+^ (memory) T-cells and pro-inflammatory CD4^+^IFN-γ^+^ (Th1) T-cells were associated with greater CCA IMT in the Multi-Ethnic Study of Atherosclerosis (MESA)^11,17^. Additionally, the treatment with an angiotensin II blocker and statin (telmisartan and rosuvastatin) elicited a coupled reduction in CCA IMT and pro-inflammatory T-cells CD4^+^IL-17A^+^ (Th17), and an increase in anti-inflammatory CD4^+^ regulatory T-cells^18^. Pro-inflammatory, CD8^+^CD57^+^ and CD8^+^CD28^-^ (differentiated/immunosenescent) have been associated with increased T-PWV^13,14^, and multiple T-cell subpopulations have also been related to elevated blood pressure or blood pressure regulation^19–21^. While results from MESA have shown that load-dependent stiffening is elevated in adults with hypertension^10^, it is not clear whether T-cells are associated with this process. To date, no studies have investigated the relationships between T-cells and S-PWV and LD-PWV arterial stiffness in large human cohorts.

The purpose of this study was to evaluate the cross-sectional relationships between T-cell populations and aspects of arterial stiffness in a large multi-ethnic, community-based cohort. We hypothesized that a higher proportion of pro-inflammatory T-cells [e.g., Th1, Th17, and CD4^+^CD28^-^CD57^+^ and CD8^+^CD28^-^ CD57^+^] would be associated with higher total pulse wave velocity (T-PWV), LD-PWV, and S-PWV, and that higher proportions of CD4^+^CD25^+^CD127^-^ (T_REG_) would be associated with lower T-PWV, LD-PWV, and S-PWV. In exploratory analyses, we evaluated associations of twenty-five additional immune cell subsets available in the MESA study with baseline T-PWV, LD-PWV, and S-PWV. In all analyses we tested whether age, sex, or race/ethnicity modified the associations.

## METHODS

### Data availability statement

MESA data can be requested from the Collaborative Health Studies Coordinating Center (CHSCC) upon approval of a paper proposal using this data. The instructions are at https://www.mesa-nhlbi.org/. MESA data is also available via BIOLINCC (https://biolincc.nhlbi.nih.gov/home/). The corresponding author can be contacted for the analytic methods. The ultrasound images are held internally at MESA to protect participant privacy. Any questions or interest in the ultrasound images, immune cell data, or other study materials should be directed to MESA via the study authors or the CHSCC.

### MESA Description

The MESA is an ongoing, prospective cohort study investigating subclinical CVD, which completed recruitment in 2000-2002^22^. MESA participants were free of known CVD at baseline and were recruited from 6 US communities (Baltimore, MD, Chicago, IL, Los Angeles, CA, St. Paul, MN, New York, NY, and Forsyth County, NC). The cohort was comprised of 6814 men and women ranging from 45 to 84 years old at baseline, and was 38% White, 28% African-American, 22% Hispanic, and 12% Chinese. Written informed consent was obtained from all participants for the study protocol. The study protocol was approved by the MESA field center Institutional Review Boards.

The present analyses leveraged data from two prior ancillary studies in MESA. The current study includes a subset of MESA participants with carotid distensibility measurements (n=6,359)^10^ and immune cell phenotype data from the baseline examination (n =2,200). The cell phenotyping data were generated from two different nested case-cohort studies^23,24^. These two sequential case–cohort studies enriched for all cases of incident coronary heart disease and heart failure and a cohort random sample. These data were leveraged for a secondary cross-sectional analysis of immune cell associations with carotid stiffness. Demographic, medical history, and laboratory data for the present study were also obtained at the baseline exam^22^.

### Carotid Ultrasound Imaging and Blood Pressure Measurements

Information regarding the carotid ultrasound image analysis has been described^10^ and can be found in the Supplementary Materials. The distal right common carotid artery was measured using B-mode ultrasound. Video-loops of a longitudinal section were recorded on S-VHS videotapes using a Logiq 700 ultrasound system (General Electric Medical Systems; transducer frequency, 13 MHz). A medical digital recording device digitized the images at a high-resolution and frame rate (PACSGEAR, Pleasanton, CA). Participants rested in the supine position for 10 minutes and repeated measures of brachial blood pressure were obtained using an automated upper arm sphygmomanometer (DINAMAP; GE Medical Systems, Milwaukee, WI) before carotid ultrasound image acquisition. The MESA Carotid Ultrasound Reading Center (the University of Wisconsin Atherosclerosis Imaging Research Program, Madison, WI) reviewed and interpreted all images.

### Calculating Total, Load-Dependent, and Structural Carotid Stiffness

T-PWV, LD-PWV, and S-PWV were calculated from the carotid ultrasound images using the Bramwell-Hill equation as previously described^10^. PWV was reported as meters/s (m/s). A participant-specific exponential model was used to differentiate the structural (S) and load-dependent (LD) components of carotid artery stiffness^25^. S-PWV was calculated as carotid PWV^26^ at the reference blood pressure of 120/80 mmHg. This reference pressure was used for all participants. LD-PWV arterial stiffness was calculated as the difference between carotid T-PWV calculated at the individuals’ measured BP and the S-PWV. If load-dependent arterial stiffness is positive, then the participant’s blood pressure is greater than the reference blood pressure, and if the LD-PWV is negative, then the participant’s blood pressure is less than the reference blood pressure.

### Immune Cell Phenotyping

The immune cell isolation procedures have been described extensively^23,27^. Briefly, CPT™ tubes were drawn during the MESA baseline examination (2000–2002) to isolate and cryopreserve peripheral blood mononuclear cells (PBMCs) at −135°C. PBMCs were thawed and then either labeled with cell surface markers, or activated with phorbol myristate acetate and ionomycin in the presence of Brefeldin A to assess CD4^+^ T helper (Th) subsets (Th1, Th2, and Th17), and CD8^+^ cytotoxic (Tc) subsets (Tc1, Tc2, and Tc17)^11^ using cell surface and intracellular antibody staining. Cells were fixed in paraformaldehyde and analyzed using a MACSQuant 10 flow cytometer (Miltenyi Biotec, Germany). Compensation controls adjusted for fluorescent carryover and isotype controls were used for negative gating. MacsQuantify software (Miltenyi Biotec, Germany) was used to analyze the data. The immune cell subpopulations were reported as percentages. Natural killer, B cells, CD4, and CD8 were reported as percent of gated lymphocytes, CD4 subsets as percent of CD4^+^ cells, and CD8 subsets as percent of CD8^+^ cells. Monocytes subsets were reported as percent of CD14^+^ gated monocytes and B cell subsets as percent of CD19^+^ B cells^23,27^. The flow cytometry gating strategy is published^23^.

### Statistical Analysis

Associations of T-cells specified in primary hypotheses (CD4^+^IFN-γ^+^, CD4^+^IL-17A^+^, CD4^+^CD28^-^CD57^+^, CD8^+^CD28^-^CD57^+^, and CD4^+^CD25^+^CD127), or secondary immune cell subpopulations with measures of arterial stiffness were evaluated using weighted linear regression models. Each immune cell subpopulation in the primary and secondary analyses was considered an independent variable and arterial stiffness measures the dependent variables. Estimates are reported per 1-SD increment of immune cell subpopulation. Multiple testing was adjusted for the 5 primary exposures using Bonferroni correction, with a significance threshold of p≤ 0.01 (0.05/5). Analyses of the secondary immune cell populations were exploratory and utilized a significance threshold of p<0.05. The model was adjusted for age, sex, race/ethnicity, cell phenotyping batch, study site, education, and log-transformed cytomegalovirus (CMV) titers. We also assessed cell-by-age, cell-by-race/ethnicity, and cell-by-sex interaction terms (F test with 3 degrees of freedom), with a significance threshold set at p-interaction <0.05. All analyses were weighted using empirical sampling weights, required because of the sampling process, to represent the whole MESA population. Continuous data were reported as mean ± SD, and categorical data were presented as percentages.

## RESULTS

### Participant Descriptives

Of the 6,814 MESA participants, arterial stiffness was measured in 6,359 participants and immune cell phenotyping was performed among 2,193 participants at the baseline study examination. Of those, 1,984 participants had both arterial stiffness measures and immune cell phenotyping and were not missing key covariates. Participant characteristics and descriptive statistics are presented in Table 1.

**Table 1.**
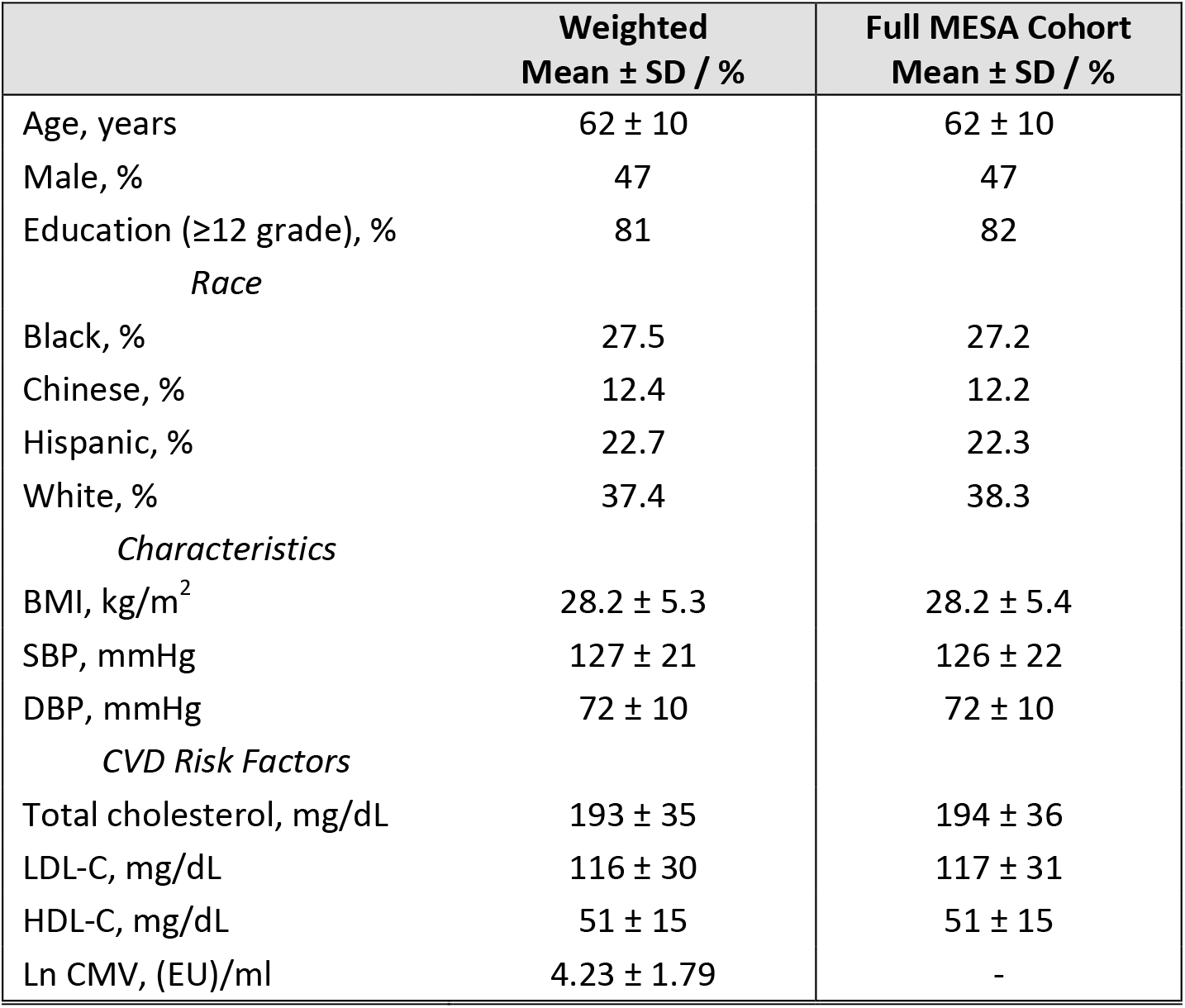
Participant Descriptives. Participant characteristics at baseline (examination 1) in MESA are presented. Data are presented as mean ± SD or percent. Abbreviations: SD, standard deviation; BMI, body mass index; SBP, systolic blood pressure; DBP, diastolic blood pressure; LDL-C, low-density lipoprotein-cholesterol; HDL-C, high-density lipoprotein-cholesterol; CVD, cardiovascular diseases; and CMV, cytomegalovirus.

### Primary Analysis

Adjusting for multiple hypothesis testing, a 1-SD higher proportion of CD4^+^CD28^-^CD57^+^ T-cells was significantly associated with higher LD-PWV (Table 2). There were no associations between the other T cell subsets specified in our a priori hypotheses including, CD4^+^IFN-γ^+^, CD4^+^IL-17A^+^, CD4^+^CD25^+^CD127^-^, or CD8^+^CD28^-^CD57^+^ T-cells with LD-PWV (Table 2). There were also no associations, of CD4^+^IL-17A^+^, CD4^+^CD25^+^CD127^-^, CD4^+^CD28^-^CD57^+^, or CD8^+^CD28^-^CD57^+^ with T-PWV or S-PWV (Table 2).

**Table 2.**
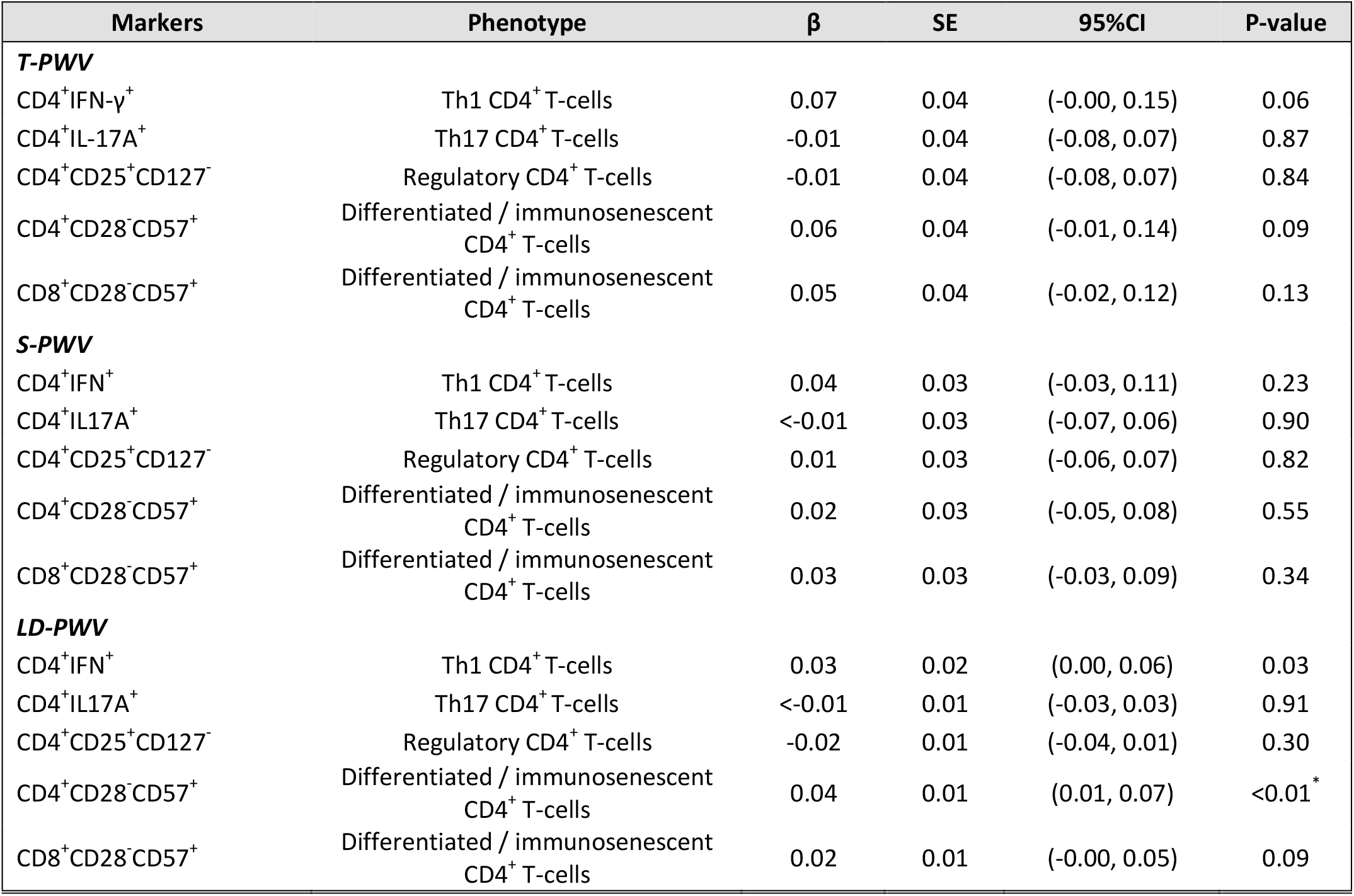
Primary analysis linear regression estimates for total (T-PWV), structural (S-PWV), and load-dependent (LD-PWV) carotid artery stiffness, weighted. Models are corrected for age, group, sex, race, site, education, and log-transformed cytomegalovirus titers. ^*^Significance for primary exposures P<0.01.

### Exploratory Analysis

#### Total Arterial Stiffness

In our exploratory analysis, without adjustment for multiple testing, each 1-SD higher CD4^+^CD45RA^+^ and CD4^+^CD38^+^ T-cells were associated with lower T-PWV; whereas, each 1-SD higher CD4^+^CD45RO^+^, CD4^+^CD28^-^CD57^+^CD45RA^+^, and CD8^+^CD57^+^ T-cells were associated with higher T-PWV (Table 3). No associations were observed for other CD4^+^ Th or CD8^+^ Tc subsets (Table 3); however, each 1-SD higher proportion of innate immune CD3^+^γδ^+^ T-cells was associated with higher T-PWV (Table 3). In analyses assessing the relationships between other immune cell populations, including monocytes, natural killer cells, and B-cells with T-PWV none were significant (Supplementary Table 1).

**Table 3.** Secondary analysis linear regression estimates for total carotid artery stiffness (T-PWV), weighted. Models are corrected for age, group, sex, race, site, education, and log-transformed cytomegalovirus titers. ^+^Significance for secondary exposures P<0.05.

| Markers | Phenotype | $\beta$ | SE | 95%CI | P-value |
| --- | --- | --- | --- | --- | --- |
| CD4<sup>+</sup> | Pan CD4<sup>+</sup> T-cells | -0.01 | 0.04 | (-0.08, 0.06) | 0.70 |
| CD4<sup>+</sup>CD45RA<sup>+</sup> | CD4<sup>+</sup> Naïve or T<sub>EMRA</sub> T-cells | -0.09 | 0.04 | (-0.16, -0.02) | 0.01<sup>+</sup> |
| CD4<sup>+</sup>CD38<sup>+</sup> | Activated CD4<sup>+</sup> T-cells | -0.14 | 0.04 | (-0.22, -0.06) | <0.01<sup>+</sup> |
| CD4<sup>+</sup>IL4<sup>+</sup> | Th2 CD4<sup>+</sup> T-cells | 0.02 | 0.04 | (-0.06, 0.09) | 0.68 |
| CD4<sup>+</sup>CD25<sup>+</sup> | T Regulatory/Activated T-cells | 0.02 | 0.04 | (-0.056, 0.096) | 0.61 |
| CD4<sup>+</sup>CD45RO<sup>+</sup> | CD4<sup>+</sup> Memory T-cells | 0.09 | 0.04 | (0.01, 0.16) | 0.02<sup>+</sup> |
| CD4<sup>+</sup>CD28<sup>-</sup> | Differentiated / immunosenescent CD4<sup>+</sup> T-cells | 0.05 | 0.04 | (-0.03, 0.12) | 0.21 |
| CD4<sup>+</sup>CD57<sup>+</sup> | Differentiated / immunosenescent CD4<sup>+</sup> T-cells | 0.03 | 0.04 | (-0.05, 0.11) | 0.44 |
| CD4<sup>+</sup>CD28<sup>-</sup>CD57<sup>+</sup>CD45RA<sup>+</sup> | CD4<sup>+</sup> T<sub>EMRA</sub> T-cells | 0.08 | 0.04 | (0.01, 0.15) | 0.03<sup>+</sup> |
| CD8<sup>+</sup> | Pan CD8<sup>+</sup> T-cells | 0.03 | 0.04 | (-0.04, 0.1) | 0.38 |
| CD8<sup>+</sup>CD45RA<sup>+</sup> | CD8<sup>+</sup> Naïve or T<sub>EMRA</sub> T-cells | -0.02 | 0.04 | (-0.09, 0.05) | 0.64 |
| CD8<sup>+</sup>CD38<sup>+</sup> | Activated CD8<sup>+</sup> T-cells | -0.05 | 0.04 | (-0.12, 0.03) | 0.23 |
| CD8<sup>+</sup>IFN-γ<sup>+</sup> | Tc1 CD8<sup>+</sup> T-cells | 0.01 | 0.04 | (-0.07, 0.09) | 0.73 |
| CD8<sup>+</sup>IL-4<sup>+</sup> | T<sub>c</sub>2 CD8<sup>+</sup> T-cells | -0.04 | 0.05 | (-0.14, 0.05) | 0.37 |
| CD8<sup>+</sup>IL-17A<sup>+</sup> | T<sub>c</sub>17 CD8<sup>+</sup> T-cells | -0.01 | 0.04 | (-0.09, 0.08) | 0.87 |
| CD8<sup>+</sup>CD45RO<sup>+</sup> | CD8<sup>+</sup> Memory T-cells | 0.04 | 0.04 | (-0.03, 0.11) | 0.27 |
| CD8<sup>+</sup>CD28<sup>-</sup> | Differentiated / immunosenescent CD8<sup>+</sup> T-cells | 0.02 | 0.04 | (-0.05, 0.09) | 0.6 |
| CD8<sup>+</sup>CD57<sup>+</sup> | Differentiated / immunosenescent CD8<sup>+</sup> T-cells | 0.10 | 0.04 | (0.03, 0.17) | <0.01<sup>+</sup> |
| CD8<sup>+</sup>CD28<sup>-</sup>CD57<sup>+</sup>CD45RA<sup>+</sup> | CD8<sup>+</sup> T<sub>EMRA</sub> T-cells | 0.04 | 0.04 | (-0.03, 0.11) | 0.27 |
| CD3<sup>+</sup>$\gamma\delta$ | $\gamma\delta$ T-cells | 0.09 | 0.03 | (0.02, 0.15) | 0.01<sup>+</sup> |

#### Structural Arterial Stiffness

Each 1-SD higher CD4^+^CD45RA^+^ and CD4^+^CD38^+^ T-cells were associated with lower S-PWV; whereas, each 1-SD higher CD8^+^CD28^-^ and CD8^+^CD28^-^CD57^+^CD45RA^+^ T-cell subpopulation was associated with higher S-PWV (Table 4). Similar to T-PWV, no associations with CD4^+^ Th or CD8^+^ Tc subsets were observed, while each 1-SD higher CD3^+^γδ^+^ T-cells was associated with higher S-PWV (Table 4). No other T-cell (Table 4) or non-T-cell subpopulations were associated with S-PWV (Supplementary Table 1).

**Table 4.** Secondary analysis linear regression estimates for structural carotid artery stiffness (S-PWV), weighted. Models are corrected for age, group, sex, race, site, education, and log-transformed cytomegalovirus titers. ^+^Significance for secondary exposures P<0.05.

| Markers | Phenotype | $\beta$ | SE | 95%CI | P-value |
| --- | --- | --- | --- | --- | --- |
| CD4<sup>+</sup> | Pan CD4<sup>+</sup> T-cells | -0.02 | 0.03 | (-0.08, 0.05) | 0.63 |
| CD4<sup>+</sup>CD45RA<sup>+</sup> | CD4<sup>+</sup> Naïve or T<sub>EMRA</sub> T-cells | -0.07 | 0.03 | (-0.13, -0.01) | 0.03<sup>+</sup> |
| CD4<sup>+</sup>CD38<sup>+</sup> | Activated CD4<sup>+</sup> T-cells | -0.11 | 0.04 | (-0.18, -0.04) | <0.01<sup>+</sup> |
| CD4<sup>+</sup>IL4<sup>+</sup> | Th2 CD4<sup>+</sup> T-cells | 0.03 | 0.03 | (-0.03, 0.10) | 0.31 |
| CD4<sup>+</sup>CD25<sup>+</sup> | T Regulatory/Activated T-cells | 0.03 | 0.03 | (-0.04, 0.09) | 0.47 |
| CD4<sup>+</sup>CD45RO<sup>+</sup> | CD4<sup>+</sup> Memory T-cells | 0.06 | 0.03 | (-0.00, 0.13) | 0.06 |
| CD4<sup>+</sup>CD28<sup>-</sup> | Differentiated / immunosenescent CD4<sup>+</sup> T-cells | 0.01 | 0.03 | (-0.06, 0.07) | 0.86 |
| CD4<sup>+</sup>CD57<sup>+</sup> | Differentiated / immunosenescent CD4<sup>+</sup> T-cells | <-0.01 | 0.03 | (-0.07, 0.07) | 0.94 |
| CD4<sup>+</sup>CD28<sup>-</sup>CD57<sup>+</sup>CD45RA<sup>+</sup> | CD4<sup>+</sup> T<sub>EMRA</sub> T-cells | 0.05 | 0.03 | (-0.02, 0.11) | 0.17 |
| CD8<sup>+</sup> | Pan CD8<sup>+</sup> T-cells | 0.04 | 0.03 | (-0.02, 0.10) | 0.23 |
| CD8<sup>+</sup>CD45RA<sup>+</sup> | CD8<sup>+</sup> Naïve or T<sub>EMRA</sub> T-cells | -0.02 | 0.03 | (-0.08, 0.04) | 0.55 |
| CD8<sup>+</sup>CD38<sup>+</sup> | Activated CD8<sup>+</sup> T-cells | -0.03 | 0.04 | (-0.10, 0.04) | 0.41 |
| CD8<sup>+</sup>IFN<sup>+</sup> | Tc1 CD8<sup>+</sup> T-cells | 0.02 | 0.04 | (-0.06, 0.09) | 0.65 |
| CD8<sup>+</sup>IL4<sup>+</sup> | T<sub>c</sub>2 CD8<sup>+</sup> T-cells | <0.01 | 0.04 | (-0.08, 0.09) | 0.92 |
| CD8<sup>+</sup>IL17A<sup>+</sup> | T<sub>c</sub>17 CD8<sup>+</sup> T-cells | -0.01 | 0.04 | (-0.09, 0.06) | 0.70 |
| CD8<sup>+</sup>CD45RO<sup>+</sup> | CD8<sup>+</sup> Memory T-cells | 0.04 | 0.03 | (-0.03, 0.10) | 0.17 |
| CD8<sup>+</sup>CD28<sup>-</sup> | Differentiated / immunosenescent CD8<sup>+</sup> T-cells | <0.01 | 0.03 | (0.02, 0.13) | 0.03<sup>+</sup> |
| CD8<sup>+</sup>CD57<sup>+</sup> | Differentiated / immunosenescent CD8<sup>+</sup> T-cells | 0.07 | 0.03 | (-0.04, 0.08) | 0.59 |
| CD8<sup>+</sup>CD28<sup>-</sup>CD57<sup>+</sup>CD45RA<sup>+</sup> | CD8<sup>+</sup> T<sub>EMRA</sub> T-cells | 0.02 | 0.03 | (-0.11, 0.01) | <0.01<sup>+</sup> |
| CD3<sup>+</sup>γδ | γδ T-cells | 0.09 | 0.03 | (0.03, 0.15) | <0.01<sup>+</sup> |

#### Load-Dependent Arterial Stiffness

Each 1-SD higher value of CD8^+^CD57^+^, CD4^+^CD28^-^, CD4^+^CD57^+^, and CD4^+^CD28^-^CD57^+^CD45RA^+^ T-cells were associated with higher LD-PWV; whereas, each 1-SD higher CD8^+^IL-4^+^ T-cell population was associated with lower LD-PWV (Table 5). No other T-cell subpopulations were associated with LD-PWV. In contrast to T-PWV and S-PWV, there was no association of CD3^+^γδ^+^ T-cells with LD-PWV (Table 5). Finally, each 1-SD higher proportion of CD3^-^CD16^+^CD56^+^ cells was associated with higher LD-PWV, with no relationships observed for any other non-T-cell subpopulation (Supplementary Table 1).

**Table 5.**
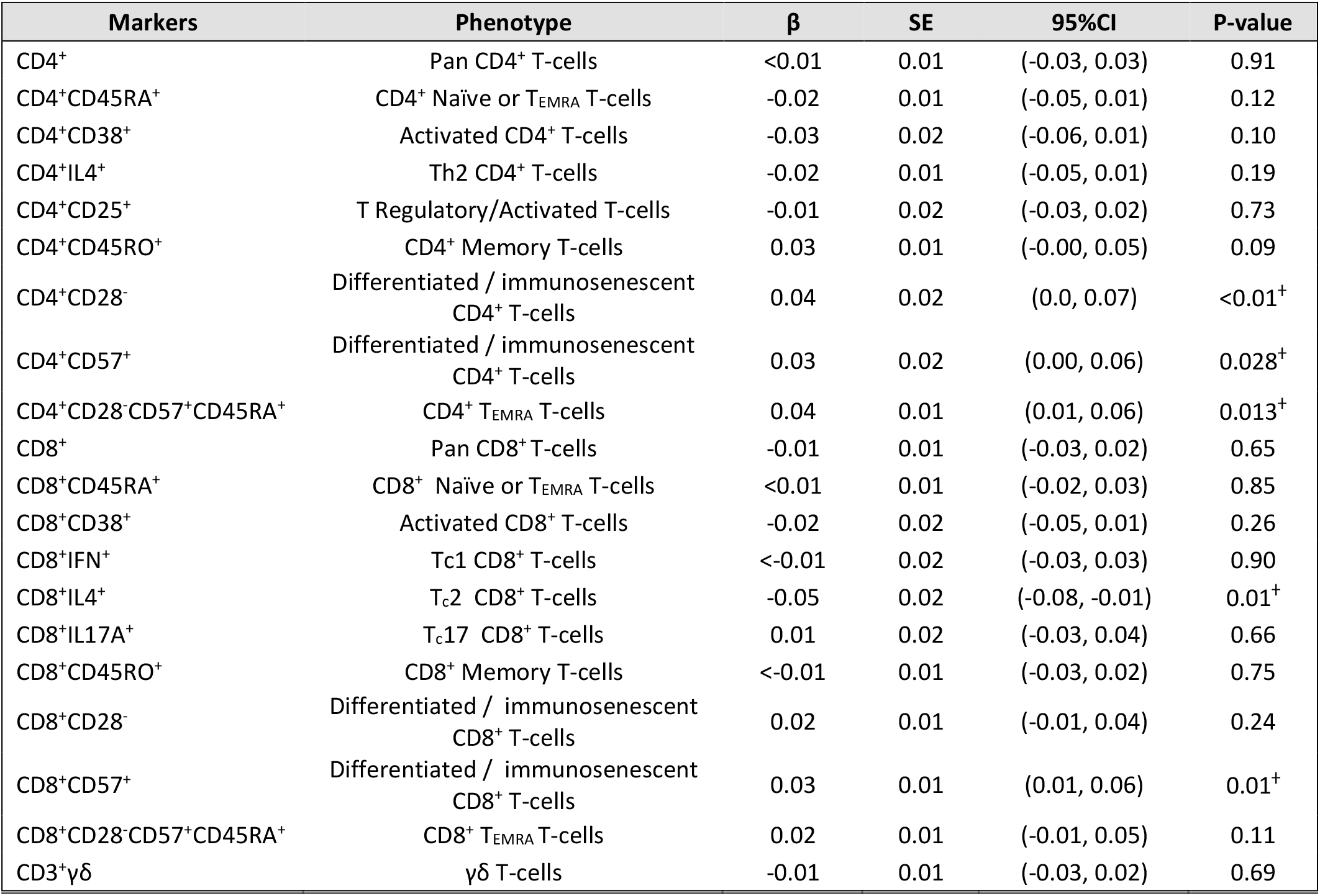
Secondary analysis linear regression estimates for load-dependent carotid artery stiffness (LD-PWV), weighted. Models are corrected for age, group, sex, race, site, education, and log-transformed cytocance for secondary exposures P<0.05.

| Markers | Phenotype | $\beta$ | SE | 95%CI | P-value |
| --- | --- | --- | --- | --- | --- |
| CD4<sup>+</sup> | Pan CD4<sup>+</sup> T-cells | <0.01 | 0.01 | (-0.03, 0.03) | 0.91 |
| CD4<sup>+</sup>CD45RA<sup>+</sup> | CD4<sup>+</sup> Naïve or T<sub>EMRA</sub> T-cells | -0.02 | 0.01 | (-0.05, 0.01) | 0.12 |
| CD4<sup>+</sup>CD38<sup>+</sup> | Activated CD4<sup>+</sup> T-cells | -0.03 | 0.02 | (-0.06, 0.01) | 0.10 |
| CD4<sup>+</sup>IL4<sup>+</sup> | Th2 CD4<sup>+</sup> T-cells | -0.02 | 0.01 | (-0.05, 0.01) | 0.19 |
| CD4<sup>+</sup>CD25<sup>+</sup> | T Regulatory/Activated T-cells | -0.01 | 0.02 | (-0.03, 0.02) | 0.73 |
| CD4<sup>+</sup>CD45RO<sup>+</sup> | CD4<sup>+</sup> Memory T-cells | 0.03 | 0.01 | (-0.00, 0.05) | 0.09 |
| CD4<sup>+</sup>CD28<sup>-</sup> | Differentiated / immunosenescent CD4<sup>+</sup> T-cells | 0.04 | 0.02 | (0.0, 0.07) | <0.01<sup>+</sup> |
| CD4<sup>+</sup>CD57<sup>+</sup> | Differentiated / immunosenescent CD4<sup>+</sup> T-cells | 0.03 | 0.02 | (0.00, 0.06) | 0.028<sup>+</sup> |
| CD4<sup>+</sup>CD28<sup>-</sup>CD57<sup>+</sup>CD45RA<sup>+</sup> | CD4<sup>+</sup> T<sub>EMRA</sub> T-cells | 0.04 | 0.01 | (0.01, 0.06) | 0.013<sup>+</sup> |
| CD8<sup>+</sup> | Pan CD8<sup>+</sup> T-cells | -0.01 | 0.01 | (-0.03, 0.02) | 0.65 |
| CD8<sup>+</sup>CD45RA<sup>+</sup> | CD8<sup>+</sup> Naïve or T<sub>EMRA</sub> T-cells | <0.01 | 0.01 | (-0.02, 0.03) | 0.85 |
| CD8<sup>+</sup>CD38<sup>+</sup> | Activated CD8<sup>+</sup> T-cells | -0.02 | 0.02 | (-0.05, 0.01) | 0.26 |
| CD8<sup>+</sup>IFN<sup>+</sup> | Tc1 CD8<sup>+</sup> T-cells | <-0.01 | 0.02 | (-0.03, 0.03) | 0.90 |
| CD8<sup>+</sup>IL4<sup>+</sup> | Tc2 CD8<sup>+</sup> T-cells | -0.05 | 0.02 | (-0.08, -0.01) | 0.01<sup>+</sup> |
| CD8<sup>+</sup>IL17A<sup>+</sup> | Tc17 CD8<sup>+</sup> T-cells | 0.01 | 0.02 | (-0.03, 0.04) | 0.66 |
| CD8<sup>+</sup>CD45RO<sup>+</sup> | CD8<sup>+</sup> Memory T-cells | <-0.01 | 0.01 | (-0.03, 0.02) | 0.75 |
| CD8<sup>+</sup>CD28<sup>-</sup> | Differentiated / immunosenescent CD8<sup>+</sup> T-cells | 0.02 | 0.01 | (-0.01, 0.04) | 0.24 |
| CD8<sup>+</sup>CD57<sup>+</sup> | Differentiated / immunosenescent CD8<sup>+</sup> T-cells | 0.03 | 0.01 | (0.01, 0.06) | 0.01<sup>+</sup> |
| CD8<sup>+</sup>CD28<sup>-</sup>CD57<sup>+</sup>CD45RA<sup>+</sup> | CD8<sup>+</sup> T<sub>EMRA</sub> T-cells | 0.02 | 0.01 | (-0.01, 0.05) | 0.11 |
| CD3<sup>+</sup>γδ | γδ T-cells | -0.01 | 0.01 | (-0.03, 0.02) | 0.69 |

#### Age Interactions

Significant interactions with age were observed for, CD4^+^IFN-γ^+^ (P_interaction_=0.01), CD4^+^CD25^+^CD127^-^ (P_interaction_<0.01), CD4^+^IL-4^+^ (P_interaction_=0.03), CD4^+^CD45RA^+^ (P_interaction_=0.01), and CD4^+^CD45RO^+^ (P_interaction_=0.03) with LD-PWV. When stratifying by age, each 1-SD higher CD4^+^CD45RA^+^ was associated with lower LD-PWV in participants older than 65 years (Supplemental Table 2). Additionally, each 1-SD higher CD4^+^IFN-γ^+^ and CD4^+^CD45RO^+^ was associated with higher LD-PWV in adults older than 65 years (Supplemental Table 2). There were no cell-by-age interactions with T-PWV or S-PWV (P>0.05).

#### Sex and Race/Ethnicity Interactions

We observed significant interactions with race/ethnicity for CD8^+^CD38^+^ (P_interaction_<0.01), CD3^+^γδ^+^ T-cells (P_interaction_=0.03) and CD3^-^CD16^+^CD56^+^ cells (P_interaction_=0.03) with T-PWV. Post-hoc testing revealed CD8^+^CD38^+^, CD3^+^γδ^+^ T-cells, and CD3^-^CD16^+^CD56^+^ cells were associated with T-PWV in Black, Hispanic, or Chinese participants but not in White participants (Supplementary Table 3). There was also a CD8^+^CD38^+^ T-cell by race/ethnicity interaction for S-PWV (P_interaction_<0.01). CD8^+^CD38^+^ T-cells were associated with higher S-PWV among Black and Hispanic participants, with no associations in White or Chinese participants. Lastly, we observed a race/ethnicity interaction between CD3^+^γδ^+^ T-cells and LD-PWV (P_interaction_=0.047) that was only significant in Hispanic participants (Supplementary Table 3). There were no sex interactions (all P_interactions_ >0.05).

## DISCUSSION

In a multi-ethnic community-based cohort of adults, we assessed the cross-sectional associations between circulating immune cell subpopulations and carotid arterial stiffness measures. In accordance with our hypotheses, higher differentiated/immunosenescent CD4^+^CD28^-^CD57^+^ T-cells, but not other primary T-cell populations, were associated with higher load-dependent carotid artery stiffness (LD-PWV). This finding suggests that CD4^+^CD28^-^CD57^+^ T-cells may be associated with risk of LD-PWV. The null findings between Th1 (CD4^+^IFN-γ^+^), Th17 (CD4^+^IL-17A^+^), T_REG_ (CD4^+^CD25^+^CD127^-^), and CD8^+^CD28^-^ CD57^+^ T-cells and LD-PWV are consistent with a previous MESA study which similarly reported no associations of these cells with blood pressure^28^. We also did not observe any significant associations in our primary analysis of Th1, Th17, CD4^+^T_REG_, or CD4^+^ and CD8^+^ CD28^-^CD57^+^ T-cells with total (T-PWV) or structural (S-PWV) carotid artery stiffness. In the exploratory analyses, that were not adjusted for multiple hypothesis testing, multiple T-cell subpopulations that are commonly associated with advancing age or repeated antigen exposure were associated with T-PWV, S-PWV, and S-PWV.

Results from the exploratory analyses are consistent with prior studies on changes in T-cell populations during aging and their roles in chronic-inflammation. Examples of these associations include a decrease in naïve T-cells and an increase in memory and differentiated/immunosenescent T-cells with advancing age^29,29–34^. While we are unable to establish whether CD4^+^CD45RA^+^ T-cells are naïve or effector memory T-cells re-expressing CD45RA (T_EMRA_) in MESA, the direction of the associations seen would suggest that this subpopulation is representative of naïve T-cells. For example, lower CD4^+^CD45RA^+^ T-cells were associated with higher T-PWV and S-PWV, whereas higher CD4^+^CD28^-^CD57^+^CD45RA^+^ (T_EMRA_) T-cells were associated with higher T-PWV and LD-PWV. Additionally, higher CD4^+^CD45RO^+^ (memory) T-cells were associated with higher T-PWV. The associations between CD4^+^CD45RA^+^ and CD4+ memory T-cells with carotid artery stiffness are generally consistent with a prior study in MESA demonstrating a similar cellular pattern was associated with measures of subclinical atherosclerosis^17^. Additionally, we observed that higher circulating levels of CD4^+^CD28^-^, CD4^+^CD57^+^, CD8^+^CD28^-^CD57^+^CD45RA^+^, CD8^+^CD57^+^, and CD8^+^CD28^-^ T-cells, which reflect various stages of differentiation and senescence, were associated with higher T-PWV, S-PWV, or LD-PWV values. Further, when stratifying by age, higher CD4^+^CD45RA^+^ T-cells were associated with lower LD-PWV, and higher Th1 and CD4^+^ memory T-cells were associated with higher LD-PWV in adults older than 65 years of age, which are generally consistent with previous findings in mid-life adults with hypertension^20,21^. Collectively, these data might suggest that loss of naïve T-cells that are generally higher in young adults, and accumulation of differentiated/immunosenescent and memory T-cells that are generally higher in older adults are associated with higher carotid artery stiffness.

We also observed an inverse relationship between CD4^+^CD38^+^ (activated) T-cells and T-PWV and S-PWV in the entire cohort. CD8^+^CD38^+^ T-cells were also inversely associated with T-PWV and S-PWV in Black participants, and positively associated with S-PWV in Hispanic participants but not among other race/ethnic groups. CD38^+^ is an enzyme involved in the metabolism of NAD^+^, an important metabolic co-factor that decreases with advancing age^35^. CD38^+^ NAD^+^ metabolism has been shown to inhibit CD4^+^ memory T-cell proliferation^36^, and may regulate the pro-inflammatory function of T-cells. We also observed a significant CD3^+^γδ T cell-by-race/ethnicity interaction term with higher T-PWV and LD-PWV in Hispanic participants. These data are consistent with previous findings in MESA showing a positive association between CD3^+^γδ T-cells and systolic blood pressure, and animal data showing the depletion of CD3^+^γδ T-cells protected against hypertension and vascular dysfunction^28,37^. Given the influence of blood pressure on LD-PWV^10^, it is plausible CD3^+^γδ T-cells are associated with LD-PWV through their relationships with blood pressure.

Our study has several limitations. First, because these data are cross-sectional, we are unable to make causal inferences, and indeed, it is possible that measures of vascular stiffness influence T-cell populations. Second, our study used cryopreserved cells which may not fully recapitulate the cells found in circulation, however data has shown that most cell types are highly correlated after cryopreservation^38^. Third, we lack more comprehensive measures of cellular senescence (e.g., P16, P21, SA-βgal) and other T-cell phenotypes (e.g., exhausted, PD1^+^ and terminally differentiated, KLRG1^+^)^39–41^ and it is controversial whether several of the phenotypes included in our study are truly senescent^31,42^. Lastly, a majority of the findings, including interactions with age and race/ethnicity, were from exploratory analyses, not corrected for multiple hypothesis testing, and future hypothesis driven studies are needed to confirm these associations.

In summary, in a population-based cohort free of clinical CVD, T-cell subpopulations that are associated with advancing age or antigen exposure (e.g., naïve, memory, and differentiated/immunosenescent T-cells) were related to greater T-PWV, S-PWV, and LD-PWV. Results may also suggest the importance of CD4^+^CD38^+^ and CD3^+^γδ T-cell subpopulations in relation to arterial stiffness in Hispanic or Black racial/ethnic groups. Although longitudinal studies are warranted to confirm these associations, these findings highlight the relationships between multiple T-cell subsets and T-PWV and its associated mechanisms (LD-PWV and S-PWV) in a large multi-ethnic cohort of older adults.

## ACKNOWLEDGMENTS

The authors thank the other investigators, the staff, and the participants of the MESA study for their valuable contributions. A full list of participating MESA investigators and institutions can be found at http://www.mesa-nhlbi.org.

